# Modeling Return of the Epidemic: Impact of Population Structure, Asymptomatic Infection, Case Importation and Personal Contacts

**DOI:** 10.1101/2020.04.26.20081109

**Authors:** Xinhua Yu

**Affiliations:** Division of Epidemiology, Biostatistics and Environmental Health, School of Public Health, University of Memphis

## Abstract

**Background:** Proactive interventions have halted the pandemic of coronavirus infected disease in some regions. However, without reaching herd immunity, the return of epidemic is possible. We investigate the impact of population structure, case importation, asymptomatic cases, and the number of contacts on a possible second wave of epidemic through mathematical modelling.

**Methods:** we built a modified Susceptible-exposed-Infectious-Removed (SEIR) model with parameters mirroring those of the COVID-19 pandemic and reported simulated characteristics of epidemics for incidence, hospitalizations and deaths under different scenarios.

**Results:** A larger percent of elderly people leads to higher number of hospitalizations, while a large percent of prior infection will effectively curb the epidemic. The number of imported cases and the speed of importation have small impact on the epidemic progression. However, a higher percent of asymptomatic cases slows the epidemic down and reduces the number of hospitalizations and deaths at the epidemic peak. Finally, reducing the number of contacts among young people alone has moderate effects on themselves, but little effects on the elderly population. However, reducing the number of contacts among elderly people alone can mitigate the epidemic significantly in both age groups, even though young people remain active within themselves.

**Conclusion:** Reducing the number of contacts among high risk populations alone can mitigate the burden of epidemic in the whole society. Interventions targeting high risk groups may be more effective in containing or mitigating the epidemic.

## Introduction

The pandemic of coronavirus infected disease (COVID-19) caused by the novel Severe Acute Respiratory Syndrome Coronavirus 2 (SARS-CoV-2) [1, 2] has significantly impacted people’s daily life and led to a global public health crisis [3]. As of April 14, 2020, there were 1,973,715 confirmed cases and 125, 910 deaths worldwide. The US accounted for 605,193 cases and 25,757 deaths (https://coronavirus.jhu.edu/map.html). To contain or mitigate the epidemic, countries and regions affected by the COVID-19 pandemic have mandated various nonpharmaceutical interventions (NPI) such as meticulous and extensive contact tracing, mass detection of virus infection, case isolation, social distancing, and closures of school and non-essential business. As a result, the pandemic was blunted in some countries, and daily new case counts are decreasing in many places [4].

However, the partially controlled or blunted epidemic leaves the source of infection and also a large pool of susceptible people in the community, posing a danger of re-surging outbreak. Two important factors may contribute to a second wave of COVID-19 pandemic. First, unlike the 2003 SARS pandemic in which mainly symptomatic cases are infectious [5], asymptomatic infection of the SARS-CoV-2 and pre-symptomatic cases can transmit the disease [6-10]. Studies have detected virus shedding in nasopharyngeal swap samples among asymptomatic cases [11]. A few case reports have shown some cluster of cases initiated by asymptomatic cases [6, 8, 10]. Researchers have postulated that asymptomatic and pre-symptomatic cases may play a significant role in sustaining the community transmission [7].

Second, government leaders have been pressed to allow people to return to normal work and life to avoid economic recession. After social activities are restored, both international and domestic travel ban will be lifted. Social and work-related gatherings are restored. Imported symptomatic and asymptomatic cases may kindle a second wave of epidemic in the community [7]. For example, despite Singapore has implemented possibly one of the most rigorous contact tracing, personal protection and social distancing measures, an unexpected surge of new cases has been observed, with newly confirmed daily cases doubled from 142 in April 7 to 287 in April 8, and still 334 in April 14 (https://www.gov.sg/article/covid-19-cases-in-singapore). As of this writing, the source of this sudden increase is still under investigation.

From the health services perspective, the number of severe cases who require medical care or die of the disease is the most important indicator of the burden of pandemic. Heath care providers, hospital beds, and intensive care units (ICU) are limited resources. One main purpose of mitigating the pandemic is to alleviate the impact of pandemic on healthcare resources. Several reports have shown that about 20% of symptomatic COVID-19 cases require hospitalizations, and of them, about 30-50% may require ICU [12-14] (also see https://gis.cdc.gov/grasp/COVIDNet/COVID19_3.html). More importantly, elderly people or people with existing chronic conditions have worse outcome than young people. For example, in the US, the mortality rate for age 50 or younger is below 1%, while the mortality rate increases to more than 10% among people aged 80 or above [15]. Finally, as demonstrated in the 2009 H1N1 flu pandemic [16], a pandemic with lower hospitalization and mortality rates has less impact on the society than those with higher hospitalization and mortality rates, though it may still have heavier impact on the economy.

Epidemic model simulation has been used extensively to estimate essential epidemic parameters, evaluate the epidemic progression and provide critical guidance to policy makers. Simulation studies based on the early epidemic data from Wuhan, China and incorporated human travel and migration information have provided more accurate picture of epidemic [17-21]. In addition, based on simulating individual behaviors under realistic societal settings, several key simulation analyses have informed the policy makers about the effectiveness of various intervention strategies to halt the epidemic [22-25]. Another simulation study updates daily about the impact of the COVID-19 pandemic on the use of health care resource and predicts the trend and peaks of health care use in the US [26] (https://covid19.healthdata.org/united-states-of-america).

In this study, we will build a modified Susceptible-Exposed-Infectious-Removed (SEIR) model [27] to simulate the COVID-19 pandemic and investigate the impact of population structure, asymptomatic cases, case importation, and the number of contacts on the epidemic progression. We will explicitly evaluate the changes of hospitalizations and mortality under various scenarios for young and elderly people. Our analysis will provide theoretical evidence for possible strategies to prepare for a second wave of epidemic.

## Method

### Modified SEIR Model

The COVID-19, like many other respiratory infectious diseases such as influenza, often has an incubation period during which the exposed persons cannot transmit the virus to others. After the incubation period, there is an infection period during which cases may or may not have symptoms but are able to infect other people. The infectivity may also vary at different time points of the infection period. As in the COVID-19 pandemic, the highest infectious points are 1-2 days around the symptom onset [28]. After the infection period, the patients are recovered or removed from the infectious pool. In addition, various controlling measures may be implemented during the epidemic, notably the case isolation, quarantine of high risk people through contact tracing, and also social distancing. All these measures will change the transmissibility of virus during a contact between an infectious person and a susceptible person. Therefore, the modified SEIR model as shown in Figure 1 is appropriate (also see the modeling framework section in supplemental documents for details). The SEIR model and its variants have been used in many previous studies for modeling the COVID-19 pandemic [23, 25]. Briefly, we divide the population into the susceptible population (S), self-quarantined susceptible people (Q), exposed but not infectious people (E), infectious compartment which includes those cases from quarantined susceptible (IQ), symptomatic cases (ID), asymptomatic cases (IU) (also those with mild symptoms, both of which are often undiagnosed or unreported), and the removed compartment which includes those hospitalized (H), recovered (R), and dead (D). The self-quarantined persons are not necessarily based on individual contact tracing but rather refer to those who are alert to any possible infection in the community and may avoid contacting with any exposed persons. They are a special susceptible people who will not infect others if they are infected.

**Figure 1:**
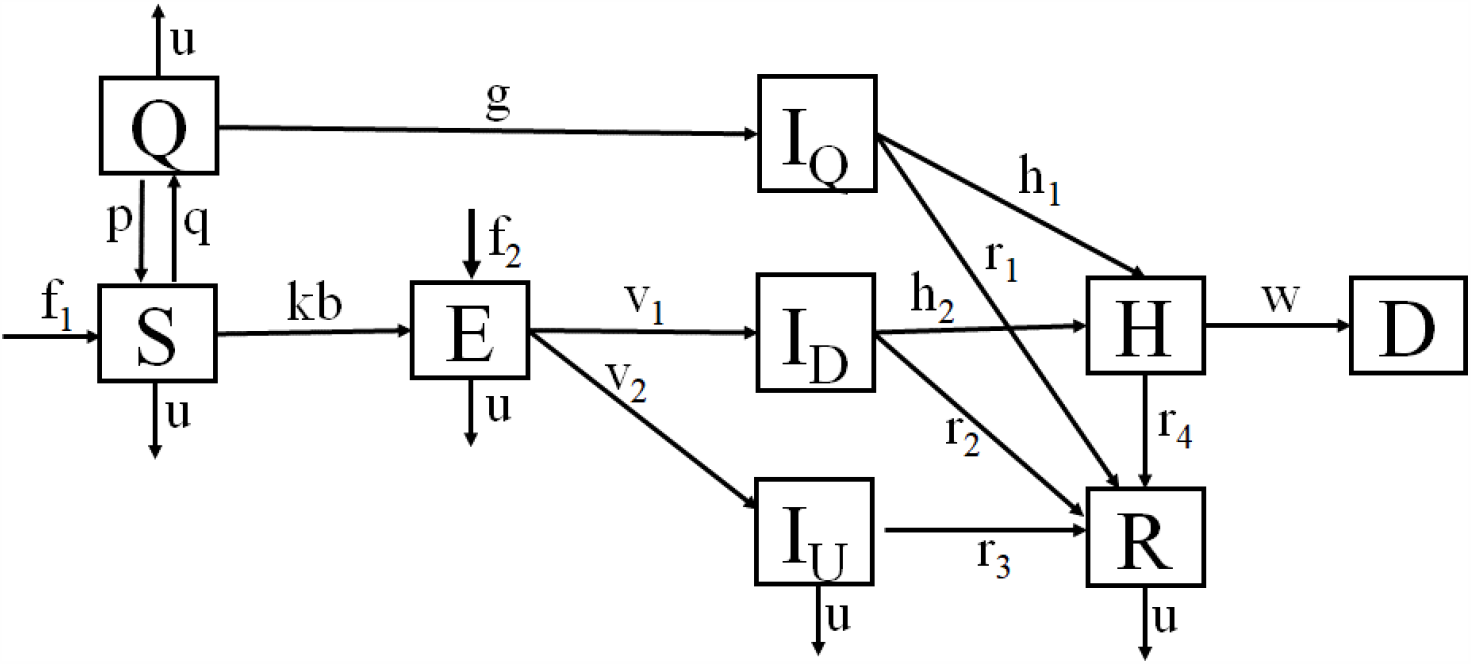
The Modified Susceptible, Exposed, Infectious, and Removed (SEIR) Model. Note: The compartments are susceptible (S), self-quarantined susceptible (Q), exposed (E), infectious (those cases from quarantined I_Q_, symptomatic cases I_D_ and asymptomatic cases I_U_), and removed (hospitalized: H, Dead: D, and Recovered: R). detailed explanations for parameters are in the text and supplemental documents

We also assume a dynamic population in which the numbers of imported susceptible persons and exposed persons (often have no symptom) are proportional (f_1_ and f_2_) to the size of total population (i.e., larger regions attract more visitors). We further assume the death rate due to other causes is a constant for all populations. For those who are symptomatic, diagnosed with the infection and hospitalized, their deaths are attributed to the infection or complications of the coronavirus infection. In addition, at the beginning of the simulation, some proportion of the population have past infection (or immunized) (P). Therefore, the total population at the time t is

N(t) = S(t) + Q(t) +E(t) + I_D_(t) + I_U_(t) + I_Q_(t) + H(t) + R(t) + P_0_. There are several other assumptions that will be discussed later and also in the supplemental document.

To account for population heterogeneity, we also apply the basic framework (Figure 1) to both young (age < 65) and elderly (age >=65) populations. The two flowcharts are connected through cross-infection due to mutual contacts. The combined flowcharts can be translated into a set of ordinary differential equations (see supplemental document). The key equations relevant to the drive of pandemic and cross-infection between two age groups are for the change of exposed people at time t (subscript y for young, and s for elderly people, with time indicator t suppressed):

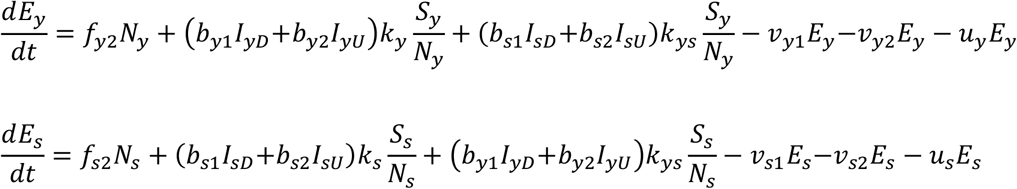

Specifically, the first equation models the exposure dynamics among young people. It includes imported exposed people (f_y2_N_y_), newly exposed people through contacting within the young people 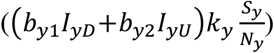 and contacting between young susceptible and infected elderly people 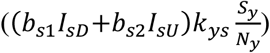. Then some percent of exposed young people become symptomatic cases (*v*_*y*1_*E*_*y*_), and some become asymptomatic cases (*v*_*y*2_*E*_*y*_). A fixed percent of exposed people will die of other diseases (*u*_*y*_*E*_*y*_). The second equation for the exposure dynamics among elderly people can be interpreted similarly.

### Model parameterization

The model involves many parameters. Their definitions, default values, and ranges are listed in the supplemental document (supp. Table 1). A few key parameters are listed in Table 1. The model parameters are set to daily rates and all the durations between various stages of disease progression are assumed with some exponential distribution. These parameters are based on observed epidemic process from China, Italy and early epidemic in the US (see references in Table 1). The key parameter, virus infectivity, is based on the basic reproduction number (R_0_), defined as the average number of secondary cases infected by an index case. Based on the basic SIR model, it can be estimated as (average contacts)*(infectivity per contact)*(serial interval). Early reports on the basic reproduction number suggested an R_0_ of 2.2, ranging from 2-3 [1]. Recent reports, however, suggested a much higher number, some as high as 5.7 [20, 29, 30]. We adopt the R_0_=2.6 as a conservative estimate[31, 32]. The number of average contacts in the population ranges 2-30 [23, 33]. We assume a moderate 10 contacts for young, 7 contacts for elderly, and 3 contacts between young and elderly people in this study. Some seniors may have more contacts than the default value due to group living or regular community gathering. They are not considered in this population level modeling. The serial interval is the average duration between the infection time point (often substituted with symptom onset time point) of the index case and the symptom onset (or diagnosis) of secondary cases. Reported serial intervals vary significantly across different studies, with an average of 5 days [17, 19, 23, 34-36]. We adopt a conservative estimate of 6 days for young and 4 days for elderly people, ranging 3-10 days. Finally, we assume an overall hospitalization rate of 10%, as commonly reported in the United States (https://www.cdc.gov/coronavirus/2019-ncov/cases-updates/). Hospital stay is 7-21 days, as most hospitalized people are elderly patients. The in-hospital mortality rate is set as 5% for young and 20% for elderly symptomatic patients, and an overall mortality of 1% and 10% for young and elderly patients, respectively [12, 14, 37-39]. The recovery duration for those who are not hospitalized is 5-20 days, typical for non-severe pneumonia.

**Table 1:**
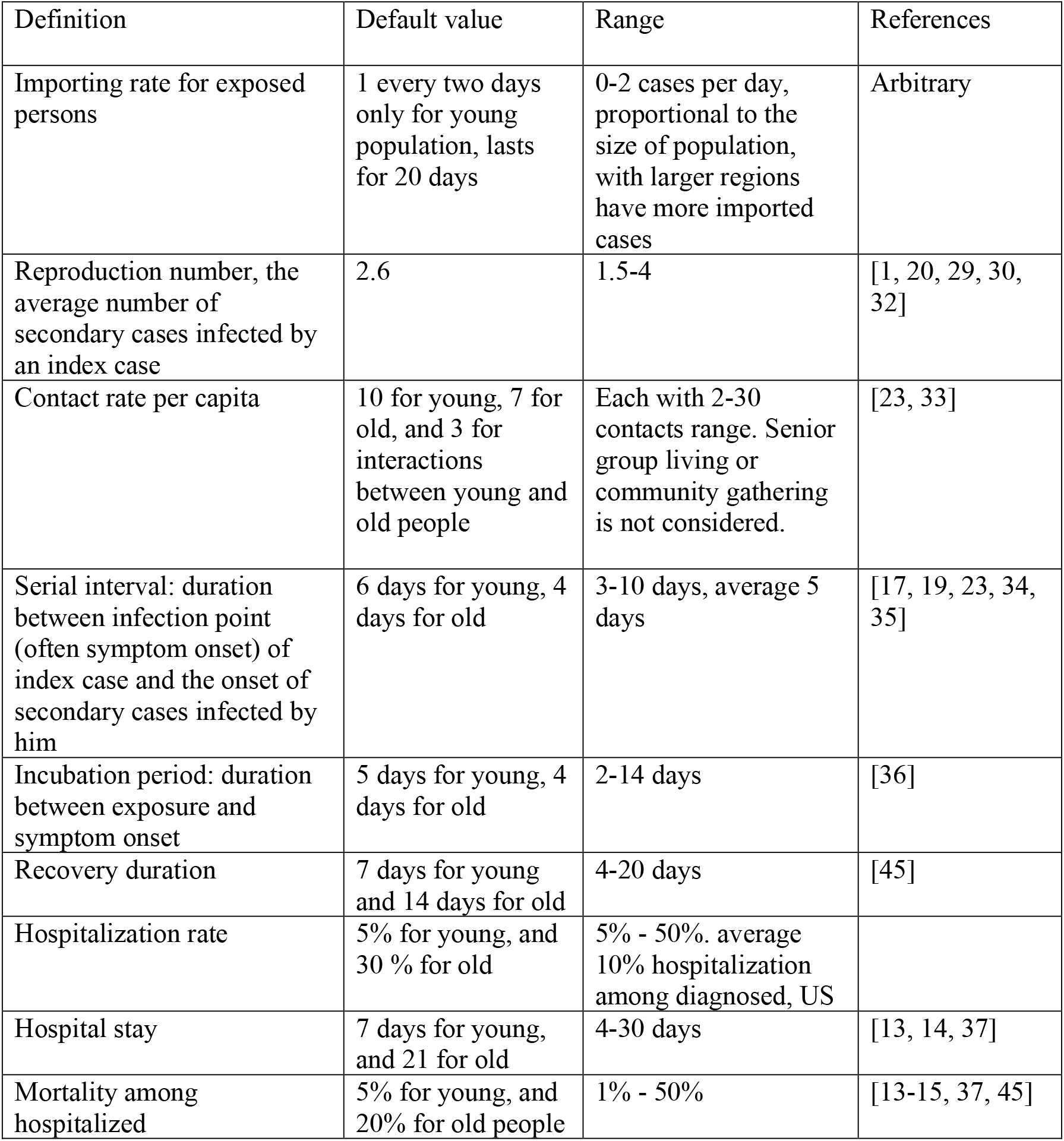
Parameters used in the epidemic models.

| Definition | Default value | Range | References |
| --- | --- | --- | --- |
| Importing rate for exposed persons | 1 every two days only for young population, lasts for 20 days | 0-2 cases per day, proportional to the size of population, with larger regions have more imported cases | Arbitrary |
| Reproduction number, the average number of secondary cases infected by an index case | 2.6 | 1.5-4 | [1, 20, 29, 30, 32] |
| Contact rate per capita | 10 for young, 7 for old, and 3 for interactions between young and old people | Each with 2-30 contacts range. Senior group living or community gathering is not considered. | [23, 33] |
| Serial interval: duration between infection point (often symptom onset) of index case and the onset of secondary cases infected by him | 6 days for young, 4 days for old | 3-10 days, average 5 days | [17, 19, 23, 34, 35] |
| Incubation period: duration between exposure and symptom onset | 5 days for young, 4 days for old | 2-14 days | [36] |
| Recovery duration | 7 days for young and 14 days for old | 4-20 days | [45] |
| Hospitalization rate | 5% for young, and 30 % for old | 5% - 50%. average 10% hospitalization among diagnosed, US |  |
| Hospital stay | 7 days for young, and 21 for old | 4-30 days | [13, 14, 37] |
| Mortality among hospitalized | 5% for young, and 20% for old people | 1% - 50% | [13-15, 37, 45] |

### Model simulation and Sensitivity analysis

The default model is set on a region with 1 million residents, consisting of 20% elderly people and 20% of total population with past infection (or immunized). There is no existing symptomatic or asymptomatic case, and no person in self-quarantine in the region. We assume only one imported young exposed case every two days for 20 days (i.e., 10 imported cases).

Analyses are performed based on the ranges of parameter estimates. We vary one parameter at a time, holding other factors at their default values. Key epidemic measures from the models are presented in tables. Multi-parameter analyses with two or more factors varying together are also performed, important findings are discussed in the text. Additional outputs are included in the supplemental materials.

The R package EpiModel is used for simulating the deterministic epidemic models[40]. The R codes for simulating the modified SEIR epidemic models are available (http://github.com/xinhuayu/returnepidemic/).

## Ethics statement

This study is deemed exempt from ethics approval as the research involves no human subjects and we use publicly available data. No informed consent is needed.

## Results

### Model calibration

Under the default model setting, all epidemic measures reflect the model parameters satisfactorily (Table 2, also refer to supplemental Table 1). That is, the resulting epidemic measures from the default model such as the disease incidence, epidemic peak, and duration of the epidemic are reasonable and mirror those reported in the literature. For example, starting with ten imported infectious persons and assuming 40% asymptomatic cases at the peak of epidemic, the epidemic reaches peak quickly within 73 days and lasts 172 days. It is ten days quicker among elderly people than among young people (Table 2). The epidemic curves for incident cases (symptomatic and asymptomatic), hospitalizations and deaths by age groups are typical (Supplemental Figure 2). The modeling results in an overall hospitalization rate of 14.1%. The in-hospital mortality rate is 5.0% for young and 20.3% for elderly people, with an overall mortality rate of 1.9%, similar to those empirical measures in the COVID-19 pandemic in the early epidemic of the US. Therefore, the default model represents the current COVID-19 pandemic sufficiently well.

**Table 2:**
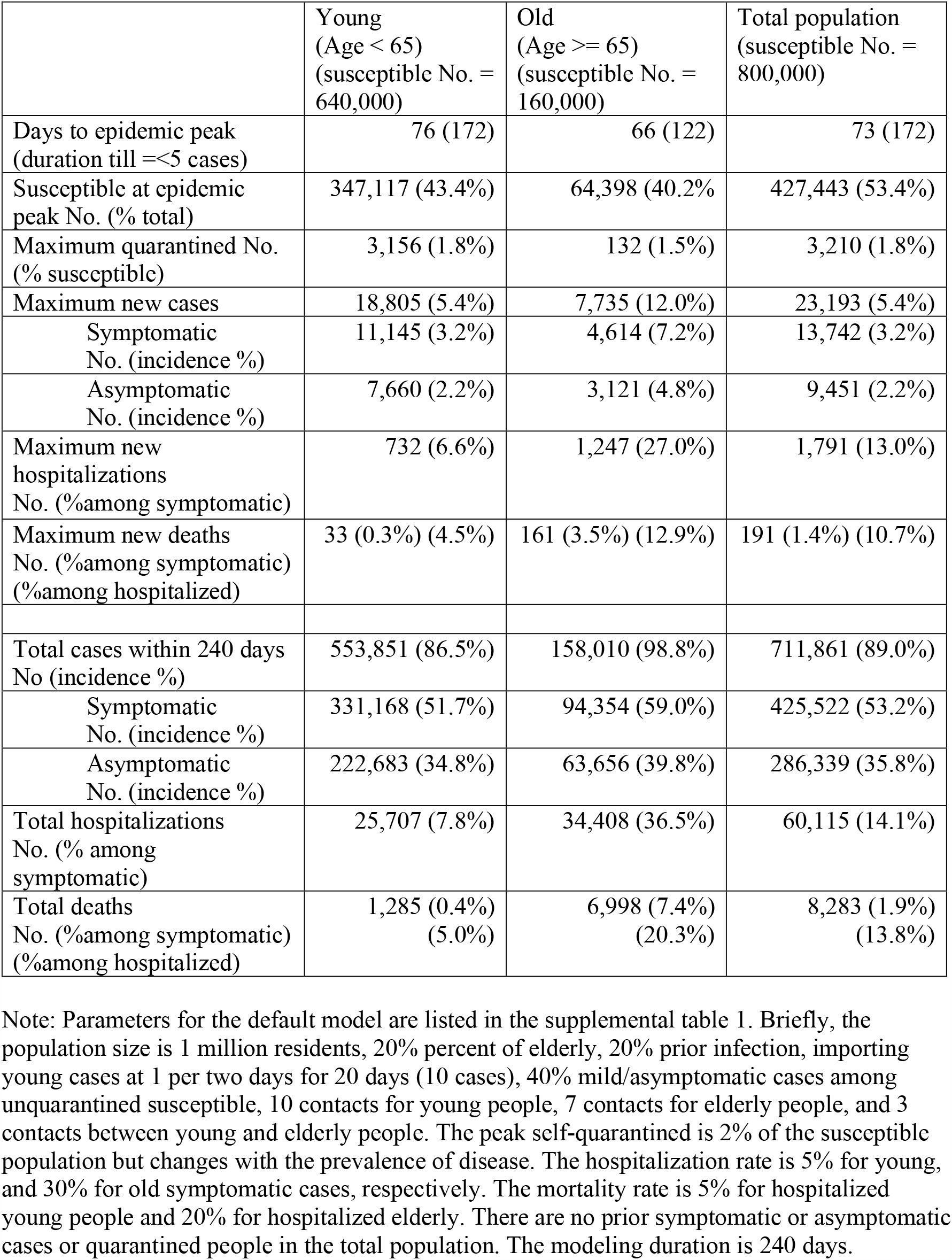
Basic epidemic measures from the simulation with default model parameterization.

### Impact of population structure

As summarized in Table 3, the size of region and a small percent change of self-quarantined susceptible people do not change the epidemic progression significantly except for the total number of cases. A smaller percent of elderly slows down the epidemic, while a much higher percent of elderly does not change the epidemic curve significantly. As expected, when over 60% people have prior infections, the epidemic takes very long to reach the peak and results in substantial fewer cases. The effects are similar in both young and elderly people (supplemental Table 2a & 2b).

**Table 3:** Impact of population structure, asymptomatic infection and importing process on the epidemic progression.

| Parameters | levels | Time to peak<br>(Duration) | Maximum at the epidemic peak |  |  |  | Total |  |
| --- | --- | --- | --- | --- | --- | --- | --- | --- |
|  |  |  | New symptomatic | New asymptomatic | New hospitalizations | New deaths | Hospitalizations | Deaths |
| Default model |  | 73 (164) | 13,742 | 9,451 | 1,791 | 191 | 60,115 | 8,283 |
| Size of region | 500,000 | 73 (164) | 6,871 | 4,726 | 896 | 95 | 30,057 | 4,141 |
|  | 100,000 | 72 (147) | 1,374 | 945 | 179 | 19 | 6,011 | 828 |
|  | 50,000 | 73 (140) | 687 | 473 | 90 | 10 | 3,006 | 414 |
| % Elderly | 10% | 77 (192) | 9,808 | 6,770 | 995 | 103 | 43,925 | 4,833 |
|  | 30% | 73 (161) | 17,108 | 11,716 | 2,500 | 273 | 75,632 | 11,716 |
|  | 40% | 73 (155) | 19,650 | 13,407 | 3,139 | 352 | 90,792 | 15,182 |
| % Prior infection | 30% | 81 (189) | 10,037 | 6,875 | 1,385 | 155 | 51,025 | 7,147 |
|  | 50% | 110 (240+) | 4,070 | 2,757 | 651 | 83 | 31,940 | 4,744 |
|  | 60% | 141 (240+) | 1,953 | 1,312 | 345 | 48 | 21,676 | 3,356 |
| % Self-quarantined | 0.5% | 73 (172) | 13,545 | 9,520 | 1,758 | 188 | 58,178 | 8,163 |
|  | 5% | 73 (171) | 14,432 | 9,239 | 1,907 | 199 | 67,121 | 8,715 |
|  | 15% | 73 (183) | 15,990 | 8,790 | 2,185 | 217 | 84,913 | 9,801 |
| % Asymptomatic cases at the peak | 10% | 54 (136) | 29,936 | 3,719 | 3,412 | 322 | 92,515 | 12,448 |
|  | 30% | 66 (160) | 17,558 | 8,501 | 2,313 | 241 | 71,646 | 10,076 |
|  | 60% | 78 (192) | 6,453 | 11,792 | 964 | 110 | 35,036 | 5,065 |
| Infectivity of<br>asymptomatic vs<br>symptomatic | 30% | 78 (185) | 12,223 | 8,388 | 1,651 | 182 | 59,076 | 8,217 |
|  | 70% | 67 (159) | 15,505 | 10,697 | 1,939 | 199 | 61,088 | 8,341 |
|  | 100% | 60 (145) | 17,997 | 12,464 | 2,135 | 210 | 62,104 | 8,397 |
| Imported young<br>cases (per day) | 0.25 | 76 (175) | 13,740 | 9,454 | 1,789 | 191 | 60,091 | 8,279 |
|  | 1 | 69 (168) | 13,760 | 9,467 | 1,793 | 191 | 60,139 | 8,287 |
|  | 2 | 65 (165) | 13,760 | 9,470 | 1,794 | 191 | 60,164 | 8,291 |
| Importing<br>duration (days) | 5 | 75 (175) | 13,730 | 9,450 | 1,790 | 190 | 60,095 | 8,280 |
|  | 10 | 73 (173) | 13,739 | 9,455 | 1,790 | 191 | 60,109 | 8,282 |
|  | 30 | 72 (172) | 13,738 | 9,455 | 1,791 | 191 | 60,116 | 8,283 |

### Impact of asymptomatic cases

Both the percent and infectivity of asymptomatic cases were investigated (Table 3). An increase of the percent of asymptomatic cases from 10% to 30% postpones the epidemic peak by 12 days due to less infectivity of asymptomatic cases, and results in significantly fewer hospitalizations and deaths. On the other hand, a higher infectivity of asymptomatic cases (e.g., 100% of symptomatic cases) results in a fast developing and narrow epidemic curve which reaches the peak within 60 days. There are more hospitalizations and deaths at the epidemic peak compared with the default model, both assumed 40% asymptomatic cases. In addition, a change of the percent of asymptomatic cases among elderly people leads to larger changes in hospitalizations and deaths than that of young people (Supplemental Table 2a & 2b). For example, comparing 60% with 40% asymptomatic cases, the total hospitalizations are reduced only by half among elderly people, while it is a two third decrease among young people. Furthermore, when the effects of the percent and infectivity of asymptomatic cases are combined, for example, in a low risk epidemic with 60% asymptomatic cases but with a lower (30%) infectivity, the epidemic reaches its peak slower for both young and elderly people with peak hospitalizations almost half of the default model (assuming 40% asymptomatic cases and 50% infectivity) (supplemental Figure 3).

### Impact of case importing process

This epidemic model is initiated by imported infectious persons (may be asymptomatic or pre-symptomatic cases). The number of imported cases is in absolute sense, regardless of the size of population. A daily arrival of two infectious people speeds up the epidemic by 8 days compared with one case every two days in the default model (Table 3). The magnitudes of epidemic are similar between different importation scenarios. In addition, a longer importing duration shifts the epidemic only slightly. Finally, if we assume all the imported cases are asymptomatic cases, the epidemic curves are not significantly different from that of default model (supplemental Figure 4).

### Impact of the number of contacts

The number of contacts affect the epidemic curves in a complicate way (Figure 2 for hospitalizations, and supplemental Figure 5 and 6 for incidence and deaths). Limiting the number of contacts only among young people changes the course of epidemic among themselves moderately (Figure 2a). It has little impact on the epidemic curves among elderly people, and subsequently does not change the burden of overall hospitalizations, as elderly people are more likely to be hospitalized or die than young people. On the other hand, limiting contacts among elderly people alone not only changes the epidemic curves among themselves, but also significantly affects those of young people (Figure 2b and 2c), even though young people maintain a lot of contacts within themselves. For example, when limiting 3 contacts among elderly people, 10 contacts among young people, and 3 contacts between young and elderly people (Figure 2c), at the peak of epidemic, the maximum number of hospitalizations is 618 for young people and 882 for elderly people. At the end of epidemic, the total hospitalizations are 25,358 and 33,741, and total deaths are 1,268 and 6,858 for young and elderly people, respectively, all significantly lower than those of default model (Table 2). The times to the epidemic peaks are also postponed in both curves. When both young and elderly people reduce contacts to 3 per day, such as under the stay-at-home rule, the epidemic curves on hospitalizations are significantly mitigated in both age groups (Figure 2d).

**Figure 2:**
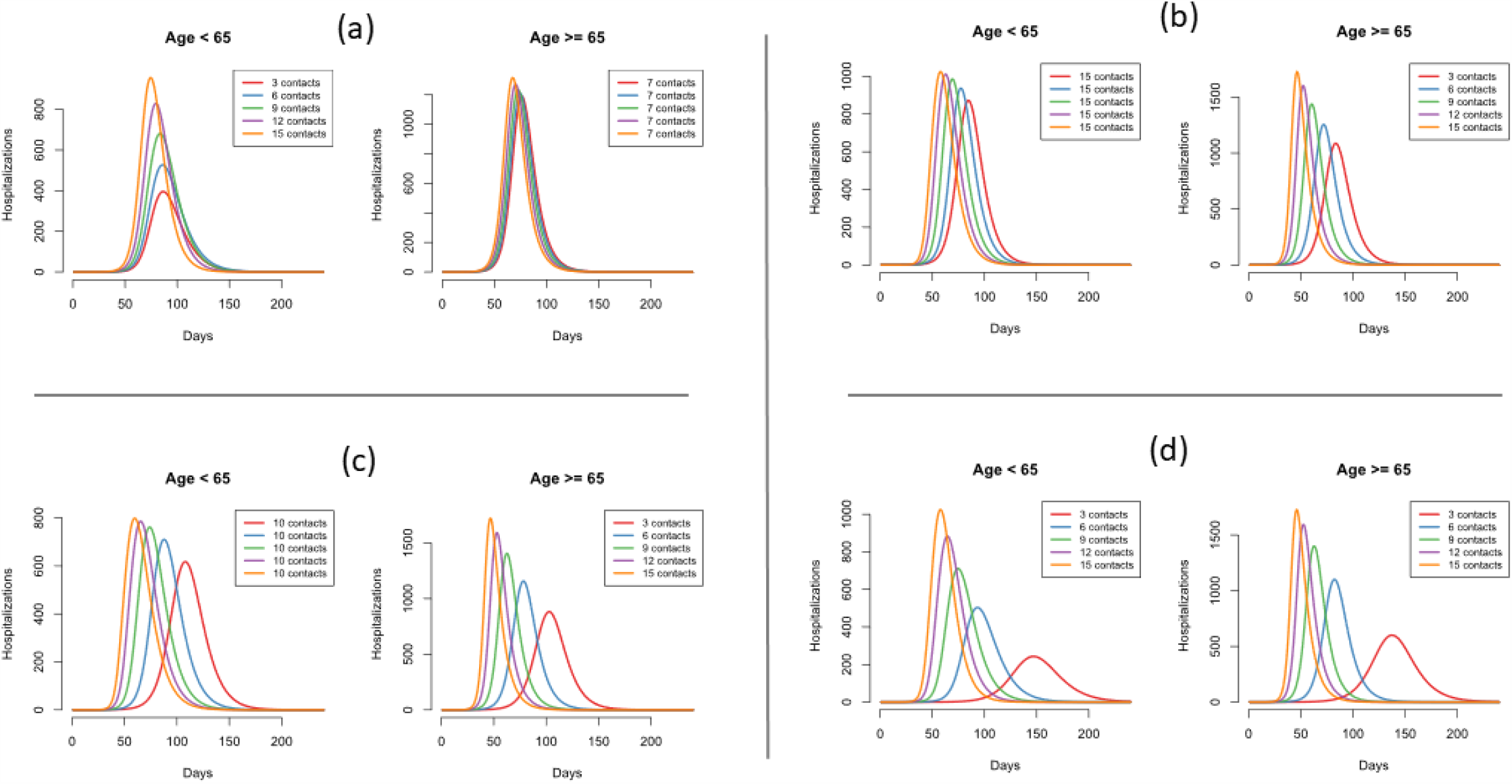
Impact of the number of contacts on the epidemic curves of hospitalizations among young and elderly people. Note: the default model includes 40% asymptomatic cases and 50% infectivity of asymptomatic cases

Finally, we consider two extreme scenarios: 1) high risk scenario: assuming one imported case per day continuously throughout the epidemic, 30% asymptomatic cases at the epidemic peak, and the same infectivity between symptomatic and asymptomatic cases; 2) low risk scenario: assuming one imported case every two days for twenty days, 60% asymptomatic cases, and asymptomatic cases have only 30% infectivity of symptomatic cases. In both scenarios, limiting contacts among elderly people alone still has significant impact on hospitalizations in both age groups, and a larger relative difference in the low risk scenario than high risk scenario (Figure 3 and supplemental Figure 7).

**Figure 3:**
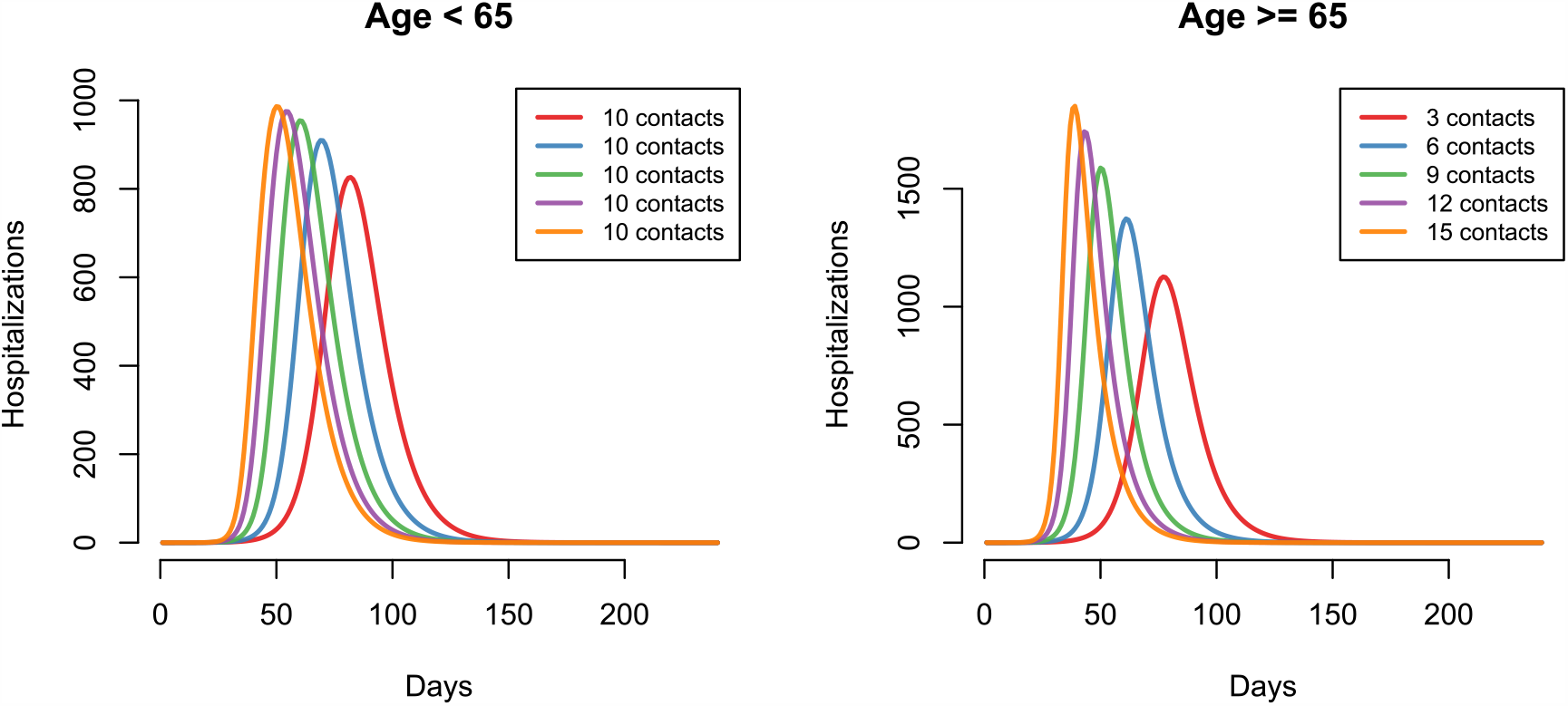
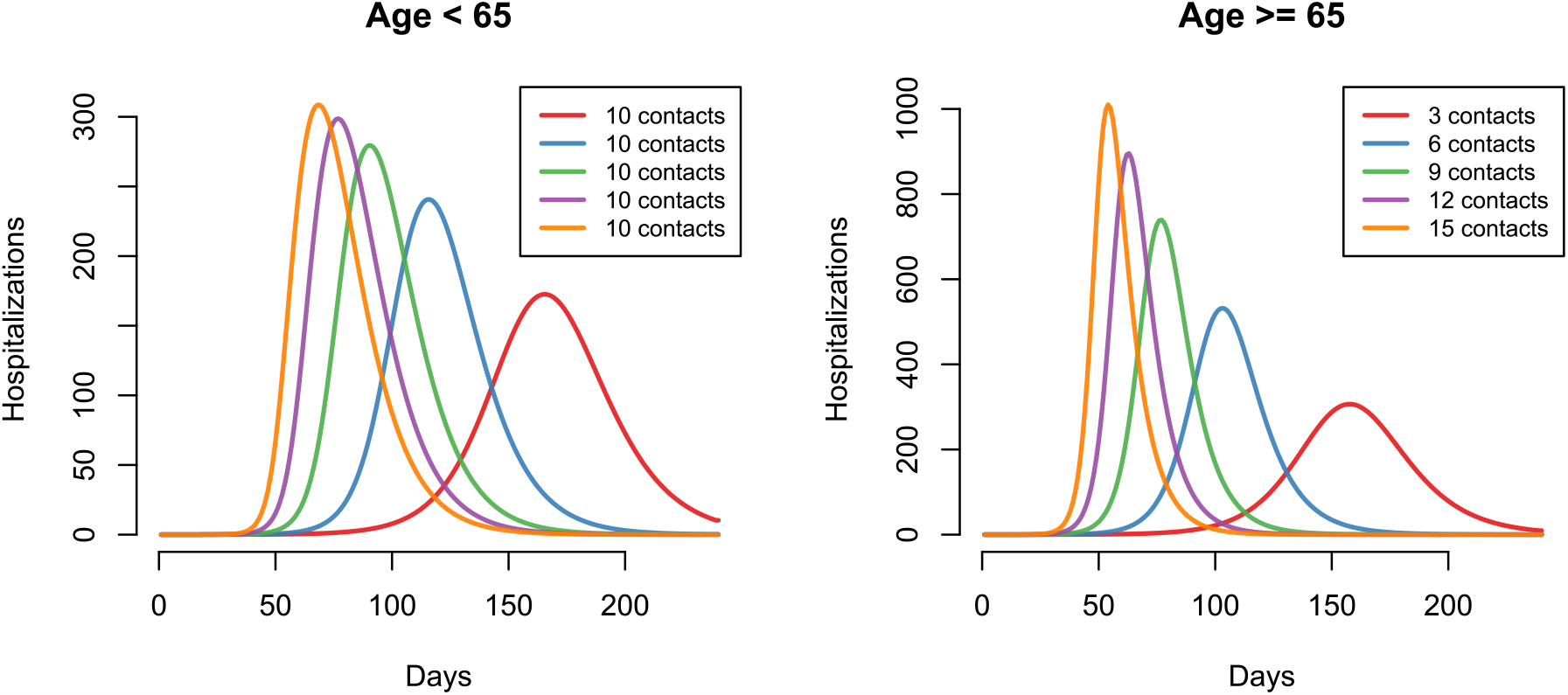
Impact of the number of contacts on the epidemic curves of hospitalizations for two scenarios. (a) High risk scenario: continuously one imported case per day, 30% asymptomatic, same infectivity between asymptomatic and symptomatic cases. (b) Low risk scenario: one imported case every two days for twenty days, 60% asymptomatic, with 30% infectivity as symptomatic cases

## Discussion

We have created a modified SEIR model and parameterized it with estimates from the current COVID-19 pandemic to investigate the impact of various parameters on the possible second wave of pandemic. Our scenario analyses suggest that unless the population have reached herd immunity (e.g., over 60% people are immune to the disease, assuming a reproduction number of 2.6 [31, 32]), a small number of imported cases within a short duration may rekindle the epidemic among the large susceptible pool. Despite young imported cases and only 3 contacts between young and elderly people, infection can quickly transmit into elderly people. Interestingly, our model demonstrates that reducing the number of contacts among elderly people alone can not only slowdown the epidemic but also reduce the magnitude of the epidemic among both young and elderly people. This spillover effect implies that interventions targeting high risk groups such as elderly population can have a larger impact on the whole society, without significantly disturbing the working life among young people.

Our findings are consistent with that of prior simulation studies and real life experience in several Asian countries [1, 17, 18, 22, 24, 26]. Many countries have implemented more proactive interventions such as school closing and stay at home rules, which have mitigated the epidemic in many regions.

One of the most effective strategies to curb an epidemic is to reduce personal interactions through social distancing and prohibiting large gatherings [23, 33, 41]. Our simulations have shown that reducing the number of contacts among young people alone does not affect the burden of epidemic significantly in terms of hospitalizations and deaths due to COVID-19. On the other hand, reducing the number of contacts among elderly people alone not only mitigates the impact of epidemic among themselves, but also changes the course of epidemic among young people, despite they still remain very active. Elderly people often have weaker immune system and multiple underlying chronic conditions. If infected with the virus, they are more likely to be symptomatic and more infectious because their bodies are not able to eliminate virus effectively. Therefore, during a pandemic such as COVID-19, elderly people are more likely to be hospitalized and die from the complications of infection. Thus, protecting elderly people not only decreases their risk of being infected but also reduces the burden of epidemic in the whole society.

With growing availability of detection kits during the COVID-19 pandemic, more asymptomatic or mild symptomatic cases are identified. Ultimately, an optimal view is asymptomatic cases may account for 60% of infections. However, recent research and case reports have confirmed that asymptomatic or pre-symptomatic cases can shed enough quantify of virus to be infectious [6-9, 11]. Furthermore, if the infectivity of asymptomatic cases is similar to that of symptomatic cases, a faster epidemic will occur. Despite more asymptomatic cases at the peak of epidemic, there are also significantly more hospitalizations and deaths, which may overwhelm the health care system. With a lower infectivity (30% infectivity) among asymptomatic cases, the epidemic reaches its peak later and results in half of hospitalizations at the peak compared with the default model (Supplemental Figure 3). In addition, a closely related issue is case importation [25].

Imported cases are often pre-symptomatic, asymptomatic or with mild symptoms. They seed of a second outbreak, even with just a few cases. Therefore, proactively identifying asymptomatic cases, isolating them and tracing their contacts thereafter will prevent the occurrence of an epidemic [22].

Our study has some strengths. We have devised a modified SEIR model to incorporate both symptomatic and asymptomatic cases. We emphasize population heterogeneity such as age structure in the model. We include a self-quarantined group who will not infect other people if they are infected with the virus. Naturally, these settings can be extended to represent other high risk or special groups with revised parameters. In addition, we separate hospitalization and death from other removed compartments to explicitly estimate the impact of an epidemic on hospitalizations and deaths. From the health impact point of view, severe cases that lead to hospitalizations and deaths are more important than mild cases, as demonstrated in the 2009 H1N1 pandemic [16]. Furthermore, we explored a few key determinants of epidemic explicitly, leading to many insights on epidemic prevention strategies.

There are a few limitations in our study. As inherent in all modeling studies, simulation interpretations are heavily dependent on model assumptions and parameter estimations. Our epidemic model is a population model. Although we take account of population heterogeneity such as age in the current model, our age group is overly broad. A more detailed age grouping scheme, including children, young adults, middle age group, and elderly, may reflect the age-specific epidemic more realistically. Other factors may also be included, and additional compartments such as pre-symptomatic stage may be modeled. However, more sophisticated models require more assumptions and may not necessarily provide more insights about the epidemic process. Instead, in this study, in addition to ensuring the mathematical correctness of the models, we prioritize the epidemiological concepts and clinical relevance in setting up the models rather than model complexity. Nevertheless, our findings do not intend to provide definitive advice to design a new policy but rather gain insights of the epidemic process and provide theoretical support for a possibly more effective prevention strategy based on approaches targeting high risk populations.

In addition, we assume random mixing within and between age groups. As a population model, we cannot assess the impact of individual behaviors such as the way of reducing contacts, social distancing and travelling. Furthermore, it ignores clustering within the population such as senior group living, community gatherings (e.g., churches, community centers), worksites and schools. These clusters are hotbeds for superspreading events which may lead to a sudden increase of new cases and overwhelm the healthcare system unexpectedly. Furthermore, the quarantine compartment in the model is not contact tracing based. Modeling contact tracing based quarantine is more relevant to public health interventions[42]. Therefore, the goal of our future research is to exploring the effect of these factors with stochastic simulations of individual behavior[24, 43] and network analysis[44]. Additionally, our model is set on a mid-size region with 1 million residents. We do not intend to model an pandemic, as all prevention strategies are ultimately local.

Our study only examines a small subset of scenarios during the epidemic. Multi-interventions and more stringent controlling measures are more effective in mitigating a pandemic but are likely less sustainable in the long run. After the initial epidemic ends, society will return to normal, and only one or two most effective interventions such as social distancing may be practiced, often partially. Thus, one parameter analysis under various scenarios is important for evaluating the probability of a second epidemic.

Finally, one critical issue in preparing for and preventing a second wave of pandemic is vaccinating the susceptible people, which is the most effective way to protect people, especially among high risk populations such as elderly people. However, there are several questions that should be addressed: a) when will the vaccine be available? A timely vaccine will protect most of the susceptible people. But despite the accelerated development of hundreds of projects worldwide, effective vaccines may still be six to eight months away; b) what percent of population will receive the vaccine? Not all people will accept the vaccination for various reasons including health conditions, allergic reactions, refusal, religious beliefs and ideologies. An insufficient coverage of vaccination may still leave a large pool of the susceptible, resulting in a possible smaller second wave of outbreak; c) how fast will the vaccine be distributed? During the pandemic, inefficient logistic may impede the vaccine distribution. Furthermore, vulnerable populations such as elderly people, people living in rural areas, people without good insurance, and some racial/ethnical groups, may receive the vaccine later than other populations, resulting in some lingering epidemics in these populations locally; and d) how effective will the vaccine be? Based on the effectiveness of the seasonal influenza vaccine, the effectiveness of the flu vaccine may only have 30-60% of protection (https://www.cdc.gov/flu/vaccines-work/effectiveness-studies.htm). In addition, virus may mutate over time, especially under the evolutionary pressure of vaccination, new strains may lose the targets of the vaccine, causing the vaccine to be ineffective. Even if the virus does not mutate over time, individual variations in the vaccine responses should not be neglected. In addition, the neutralizing effect of the vaccine has not been established, and a short-term immunity against the novel SARS-CoV-2 will undermine the purposes of vaccination. Currently we are conducting further analyses to examine these issues.

In summary, with a modified SEIR model, we have demonstrated that simple intervention strategies such as reducing the number of contacts through social distancing among elderly people alone will not only reduce the risk of infection and alleviate disease burden among themselves, but also mitigate the impact of a second epidemic in the whole society. Interventions targeting high risk groups may be more effective in containing or mitigating the pandemic.

## Data Availability

available upon request

## Acknowledgments

The author wishes to acknowledge the support from FedEx Institute of Technology at the University of Memphis for the data science cluster seed grant fund. The funder plays no role in design, analysis and report the current study. The author also wants to thank two anonymous reviewers for their insightful comments and suggestions that significantly improved the manuscript.

